# EpiAlert: A Transparent CUSUM-EWMA Framework for Early Warning of Unusual Influenza-Like Illness Surveillance Activity

**DOI:** 10.64898/2026.10.09.26365189

**Authors:** M Varshith Reddy, N Sai Varshith Reddy, J Vijay Ram, K Nandini

## Abstract

Automated surveillance systems can prove to be extremely useful in detecting outbreaks of respiratory diseases that need further epidemiological investigation. In order for the system to be effective, careful attention has to be paid to the baseline used, absence of the observations, and the use of proper statistical methods. As a solution, the paper describes EpiAlert, a weekly surveillance system based on CDC FluView ILINet. The system is able to transform the number of ILI visits and the total number of visits into a ILI prevalence for the region, create the first reference from the three previous years using a +/-2-week interval, transform the rate value, and apply two different detectors - CUSUM and EWMA. CUSUM is set at k=0.5 and h=5, whereas EWMA uses λ=0.2 and H=2.7 values. Among the 55 districts, the system gives 45,138 pieces of information about the disease by transforming 44,297 pieces of data into alerts. It is found that there are 2,820 alerts in total (out of which 2,711 alerts used CUSUM, 402 used EWMA, and 293 used both detectors). Additionally, the experiments show that the sensitivity of the device depends greatly on the detector. The latest statistics can be tracked in the report for week 37 of 2026 that recently concluded in West.

## I. INTRODUCTION

The identification of unusual respiratory illness activity in a timely manner is a key aspect of surveillance work. Weekly syndromic data are affected by seasonality, variations in the amount of reported data, missing data, etc. Additionally, the definition of this kind of surveillance does not identify the specific pathogens present. Statistical process control offers a scientific way to track the series and send an alert when the data deviate from the baseline.

The FluView system from the CDC is based on various sources of data. For instance, ILINet collects the data on outpatient visits due to ILI from various health care institutions in the US and in some jurisdictions. According to the CDC, ILI includes fever of 100°F (37.8°C) and cough and/or sore throat, and the CDC stresses the fact that ILINet is a syndromic type of surveillance, not the laboratory-confirmed one. Therefore, the increase in the indicator EpiAlert should be treated as an unusual ILI observation calling for investigation rather than as evidence of any influenza epidemic.

This creates a key distinction for EpiAlert: it was not created to determine presence of an influenza outbreak; rather, its primary function is more limited, that is, to find the region-week findings, where the ILI activity is statistically different from historical averages.

Hereby, this project tackles the methodological issue discovered earlier in disease count implementation, because rare counts often have many zeros in them. ILINet offers several advantages due to the continuous nature of the measure, and via use of a proper denominator the detecting becomes possible for the ILI rate instead of counts of rare diseases.

The main contributions of the study are: (1) the replicable process of data preparation; (2) the specific seasonal average for study region; (3) two different implementations that are accessible and can be checked; (4) approaches to handling gaps and nulls; (5) sensitivity analysis of parameters and average; and (6) the dashboard that enables one to see the output of analysis without hiding main values.

## II. RELATED WORK

Automated detection of aberrations has a long history in public health monitoring. Page pioneered the cumulative sum method for sequential detection of change, and Roberts developed the geometric/exponentially weighted moving average control chart method. Further research yielded EWMA methodology, regression-based thresholds for outbreaks, and practical aberration detection systems such as EARS. Studies show that the performance of an aberration detector depends on the baseline, seasonal effects, data properties, thresholds, and the shape of the signal over time.in the EpiAlert system to demonstrate that it utilizes meaningful and measurable options. Evaluations of epidemiological detection mechanisms have revealed that various aspects directly impact the ability to detect abnormalities [5]. A systematic review of the results of 145 studies related to disease detection indicated that statistical approaches are the most widely researched and applied, but the scientific community became more interested in non-statistical methods, such as forecasting, machine learning, and Bayesian approaches [6].

The EpiAlert system pursues the goal of developing easy-to-understand detection systems and avoids creating and using non-transparent prediction methods. Therefore, the research question is not which method is better but rather how the two complementary temporal methods behave.

## III. RESEARCH GAP AND OBJECTIVES

Implementing effective surveillance requires more than just developing an algorithm. Beyond this, a surveillance system should delineate the surveillance unit, create a suitable baseline, process the unobserved events instead of turning them into false zeros, and present evidence for every alert. Previous research on surveillance underscored the importance of evaluating the efficiency of any detection algorithms within the context of data used and signal characteristics [7], [10], [11]. EpiAlert is thus aimed at the reproducibility and interpretability of results, not the achievement of the absolute optimal detection method. The main aim of EpiAlert is to develop and evaluate the transparent statistical surveillance system for unusual ILI incidences on the level of regions answering three questions:

- How does using seasonal reference based on previous years influence the signal generation compared to the trailing rolling reference?
- What is the difference in signal frequency received through CUSUM compared to EWMA when dealing with the same standardized input?
- How sensitive are the outcomes to the parameters of the detector?

## IV. EPIALERT ARCHITECTURE

**Fig 1.**
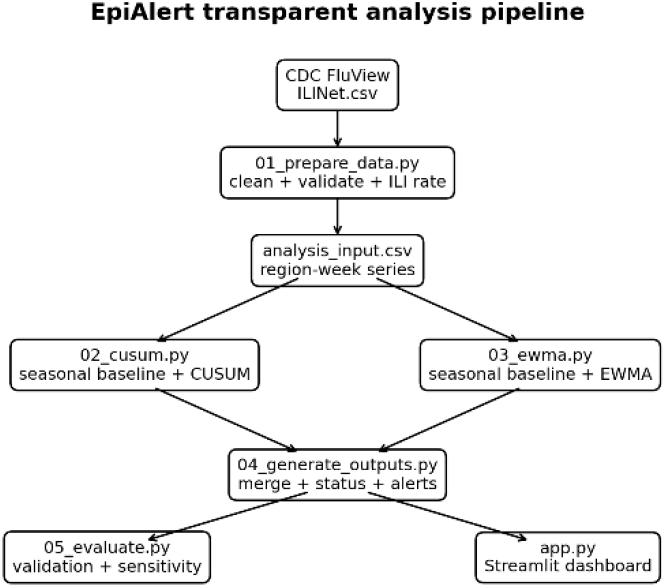
Transparent EpiAlert pipeline used in the current ILINet study.

This project is implemented as a clear chain of scripts. First, the input CDC data are read. Then, preprocessing generates an analysis table which has been validated. The calculation of CUSUM and EWMA is done in separate steps.

Finally, the generated output combines the results. The evaluation generates summary tables and figures that are displayed by the Streamlit dashboard. This is consistent with the surveillance methodology used to convert the original data into the monitored time series and the application of specific anomaly rules [13], [14]. By segregating these details, each of the detectors could be evaluated independently with the capacity to recreate the final output with similar raw data. The output table presents information such as region, year, week number, ILI counts, ILI denominator, ILI rate, statistics related to baseline, CUSUM, EWMA, detector status, alerts status, and data status.

## V. DATASET AND PREPROCESSING

The analysis employs the given CDIC FluView ILINet CSV export. The resulting unprocessed table comprises 45,138 regional-week observations encompassing 55 reporting areas from 2010 to 2026; 2026 is only preliminary until the end of the 37th week (MMWR Week 37). According to the CDC, ILINet collects the number of weekly ILI visits by the participating outpatient service providers, making it possible to present the data with a rate-based indicator because of possible variations in the volume of visits in different years that are compared.

**TABLE I.** ILINet Fields and Their Roles in EpiAlert

| Field | Role in EpiAlert |
| --- | --- |
| REGION | Independent surveillance series |
| YEAR, WEEK | Chronological ordering |
| ILITOTAL | Reported ILI visits |
| TOTAL PATIENTS | Reported patient-visit denominator |
| ILI_RATE_PCT | Primary detector input |
| NUM. OF PROVIDERS | Reporting-volume context |

The primary surveillance measure is the reported ILI visit rate:

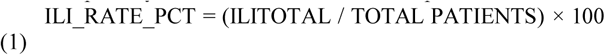

The utilization of the rate helps minimize the dependency on pure reporting numbers, but the numerator and denominator values are still available for further analysis. Thus, the sensor works with a kind of indicator in the form of a rate rather than through the use of raw ILI visit statistics [1].

The numeric fields are processed through various transformations (conversion of each field to numbers). The non-numeric data, including ‘X’, is treated in a way that it is considered to be “missing” but not as zero. If there is an explicit numeric “0”, it will remain. The data records that are missing either ILI numbers or the denominator (including non-positive) are not eligible for calculation through the detector. The raw row in the processed data retains its origin in terms of chronology.

The quality control procedures lead to the discovery of 772 missing ILITOTAL and denominator values, 999 rows with zeros, 69 cases of non-positive denominators, and zero duplicates. The total number of valid observations for the rate-based detection is 44297

## VI. SEASONAL BASELINE AND STANDARDIZATION

The prevalence of influenza-like illnesses is based on seasons, and while making reference to previous weeks only may confused the usual seasonal rise with some unpredictable happening, it is indeed obvious that seasonalities and the baseline comprise the core elements of successfully detecting outbreaks with any surveillance system. For that reason, EpiAlert applies the method of the same calendar week neighborhoods of the previous years [5], [7], [11], [14] in its operations. There must be at least 12 historical observation points available. The seasonal data is processed using log(1+x), where x is the ILI rate. This reduces the influence of very large rate peaks while preserving zero as log(1)=0.

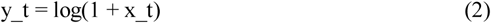

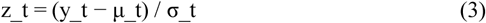

The term μ_t is used to refer to the mean while σ_t refers to the sample standard deviation. Positive and negative z-score values indicate the degree of activity with respect to historical information. The detector will not work in instances when there is no historical reference available for the given period.

## VII. CUSUM DETECTOR

CUSUM from the upper side collects evidence to ensure that standardized surveillance rate is continuously more than the reference rate. This follows sequential cumulative-sum principle as described by and discussed in prospective surveillance literature [7], [11]. Implementation:

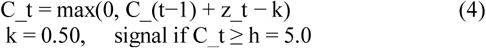

[2]

The margin divides minor positive deviations from contributing fully in terms of their magnitude to the cumulative measure. The detector is upper-sided and the negative evidence pushes the accumulator towards zero without allowing it to go negative. After the signal, the accumulator is reset to zero. This reset allows the future signals to be treated as new episodes of accumulating evidence instead of a continuous increase in the measure.

## VIII. EWMA DETECTOR

EWMA gives a different version of the temporal memory. The concept of a geometric moving average control chart was proposed by Roberts. The subsequent work formalized the properties and design considerations of EWMA. EpiAlert utilizes this recursive statistic:

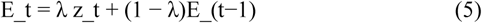

[3,4]

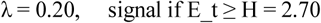

A smaller λ means that the EWMA’s historical state will have more weight and produce a smoother statistic. In comparison to the upper CUSUM, EWMA does not have to be non-negative. Hence, having a negative EWMA is valid and indicates that recent standardized activity has been generally below the seasonal benchmark. In the current implementation of the method, the EWMA state is reset after receiving a signal.

## IX. ALERT DEFINITION AND INTERPRETATION

**TABLE 2.** EpiAlert Alert Status Definitions

| Status | Definition |
| --- | --- |
| NORMAL | Neither detector crosses its upper threshold. |
| CUSUM SIGNAL | CUSUM crosses 5.0. |
| EWMA SIGNAL | EWMA crosses 2.70. |
| UNUSUAL ACTIVITY DETECTED | Both detectors signal for the same region-week. |

A signal simply means an indication of statistical investigation instead of an interpretation of it. The distinction follows syndromic-surveillance practices, where statistical signals serve the purpose of marking observations for further examination without establishing a causal link, nor confirming the presence of an outbreak. CDC warns that ILINet counts ILI visits but not laboratory-detected cases of respiratory infections and needs to be understood alongside other data.

## X. EVALUATION DESIGN

We evaluated the entire supplied historical series instead of using synthetic outbreaks to obtain the desired results. Therefore, we aimed to understand the behavior of the detectors, data quality, parameter sensitivity, baseline sensitivity, and current surveillance operations. This is intentionally a descriptive evaluation: without an independently curated outbreak truth set, standard sensitivity/specificity measures cannot be utilized. Previous studies have shown that using formal detector comparisons requires explicit reference events or controlled simulations [9],[12],[15].

A secondary rolling baseline has been constructed for the purpose of comparison. This baseline draws from the previous 26 valid values for each region instead of making use of a seasonal historical window. It is a descriptive comparison and not one that posits the superiority of one baseline over others.

In the experiments, parameter sensitivity sets CUSUM k at 0.25, 0.50, and 0.75 with h taking the value of 4, 5, and 6. EWMA λ was set at 0.10, 0.20, 0.30, and 0.40 and the thresholds at 2.5, 2.7, and 3.0. This series of experiments allowed for understanding how the alert volume changes due to the nature of the monitoring design.

## XI. RESULTS

The ultimate dataset comprised 45,138 records obtained from 55 areas over a period of 17 years. Out of these records, 44,297 were valid for detection purposes. The main seasonal detection system showed 2,820 alerts (6.25% of valid records), from which 2,711 were produced by the CUSUM detector, while 402 alerts were signaled by EWMA, and there were 293 cases signaled by both systems. The concordance shows that detectors provide different temporal responses and not identical decisions.

**Table 3.**
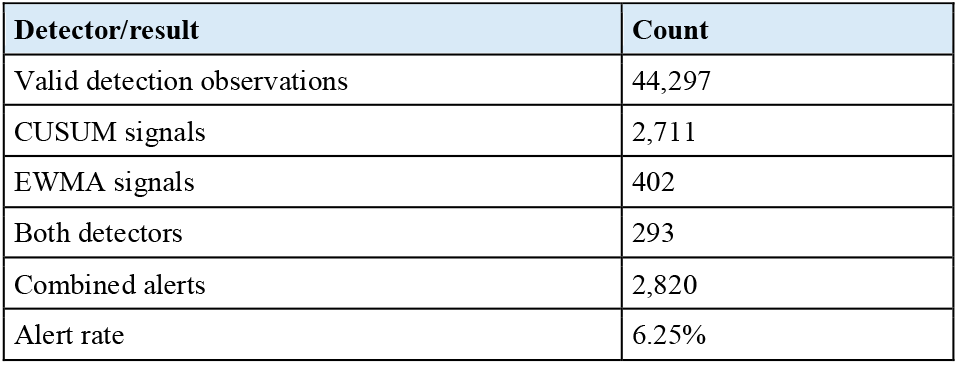
Overall EpiAlert Detection Results

| Detector/result | Count |
| --- | --- |
| Valid detection observations | 44,297 |
| CUSUM signals | 2,711 |
| EWMA signals | 402 |
| Both detectors | 293 |
| Combined alerts | 2,820 |
| Alert rate | 6.25% |

**Fig 2.**
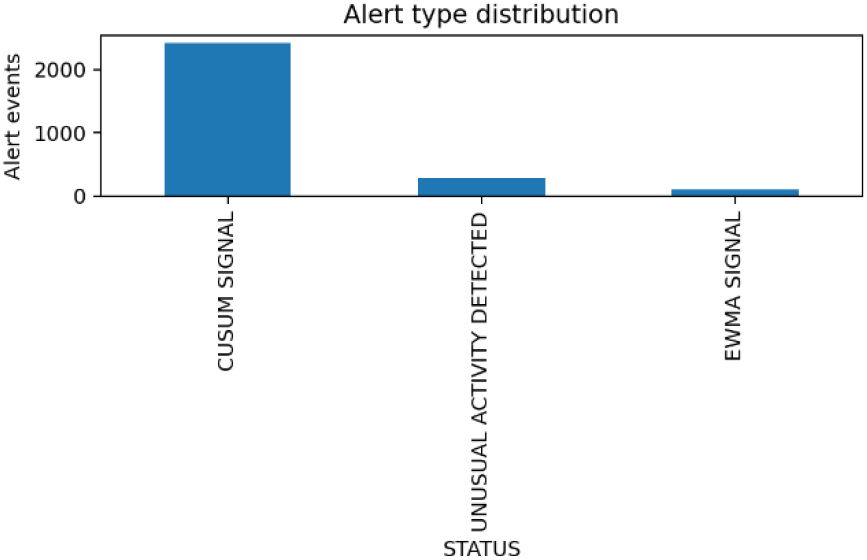
Distribution of final EpiAlert status categories.

According to the overlap diagram presented, out of 2848 total observations, 2418 corresponded only to CUSUM, 109 were only detected by the method of EWMA, and the remaining 293 were flagged using both methods. Hence, it can be concluded that CUSUM has yielded more signals relative to the parameters considered. However, this is a limitation of the detection setting selected rather than proof of the superiority of CUSUM. Published comparisons demonstrate that other algorithms respond differently to signal size, duration, background level, and number of alerts [9].

**Fig 3.**
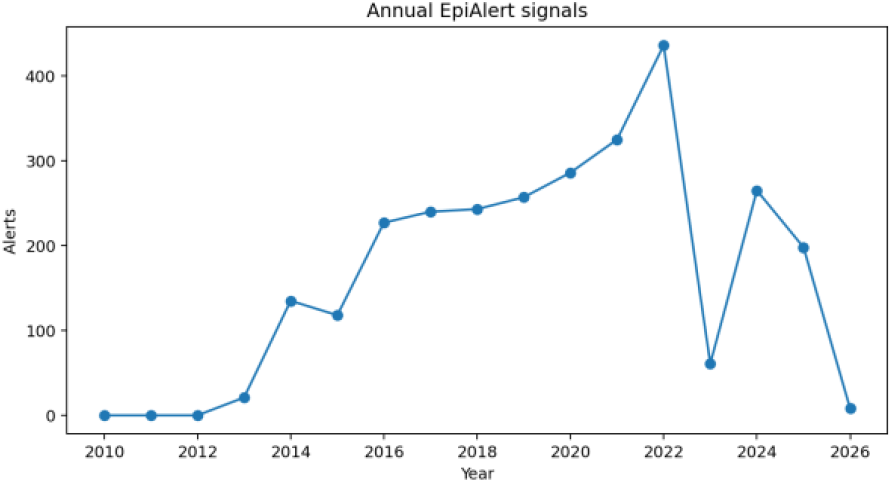
Annual EpiAlert alert observations in the supplied dataset.

The yearly sequence must not be viewed as a count of outbreaks. It is made up of the observations for the region-week level that went over chosen statistical levels. The counts of alerts are going to be increasing in the coming years, while the early years show a weak signal or none because of seasonal reference that requires three prior calendar years. The 2026 value is incomplete, and thus it should not be compared with complete years directly.

## XII. PARAMETER AND BASELINE SENSITIVITY

The findings of sensitivity analysis showed that the volume of alerts is greatly influenced by parameters of the detector. The signals for the tested grids of CUSUM varied from 1,858 to 4,070, while signals for the tested method of EWMA changed from 106 to 1,129.

**Table 4.** Parameter Sensitivity of CUSUM and EWMA Detectors

| Method | Setting | Signals |
| --- | --- | --- |
| CUSUM | $k=.25, h=4$ | 4,070 |
| CUSUM | $k=.50, h=5$ | 2,711 |
| CUSUM | $k=.75, h=6$ | 1,858 |
| EWMA | $\lambda=.10, H=2.7$ | 146 |
| EWMA | $\lambda=.20, H=2.7$ | 402 |
| EWMA | $\lambda=.40, H=2.7$ | 931 |

Such an influence of configuration is expected since it is typical of sequential methods of monitoring via charts, and hence highlights that parameters need to be reported clearly rather than using thresholds as general epidemiological reference points.

**Fig 4.**
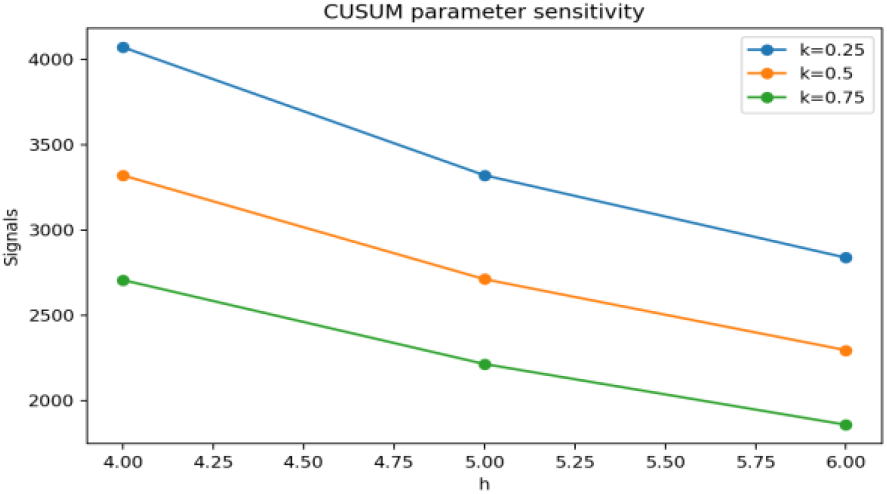
CUSUM signal sensitivity to k and h.

**Fig 5.**
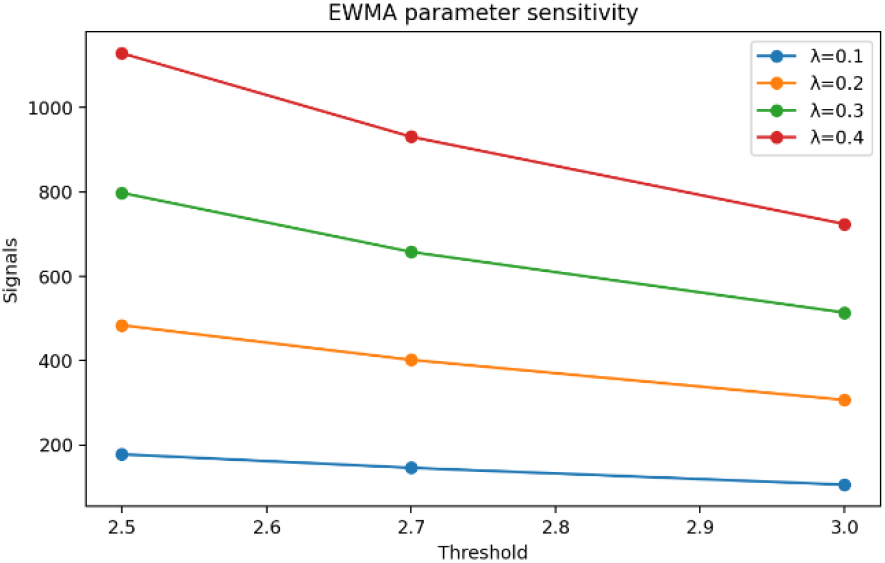
EWMA signal sensitivity to λ and threshold.

**Table 5.** Comparison of Seasonal and Rolling Baselines

| Baseline | CUSUM | EWMA | Combined |
| --- | --- | --- | --- |
| Seasonal: previous 3 years, $\pm 2$ weeks | 2,711 | 402 | 2,820 |
| Rolling: previous 26 valid observations | 3,250 | 145 | 3,314 |

The seasonal baseline produced 2,820 alerts while the alternative rolling baseline recorded 3,314. This difference reveals that the baseline is not merely an aesthetic preprocessing option since the distribution that it refers to changes the amount of signals produced. EpiAlert uses the seasonal baseline because its target variable is highly seasonal, and the method specifically compares similar calendar-week neighborhoods in different years [1], [5], [11].

## XIII. CURRENT SURVEILLANCE VIEW

The provided dataset goes well beyond MMWR Week 37 of 2026. During this week, EpiAlert made one signal, which was in West Virginia. The figure for the total number of visits noted was 370 ILI visits out of 13407 visits generating ILI rate of 2.7598%. The standardized deviation of this number was z=2.2123 and the CUSUM statistic was 6.4629 and EWMA was 1.1634 which shows that CUSUM signal was triggered and EWMA did not. This is a statistical result from the processed dataset and not the CDC classification of an outbreak.

**Fig 6.**
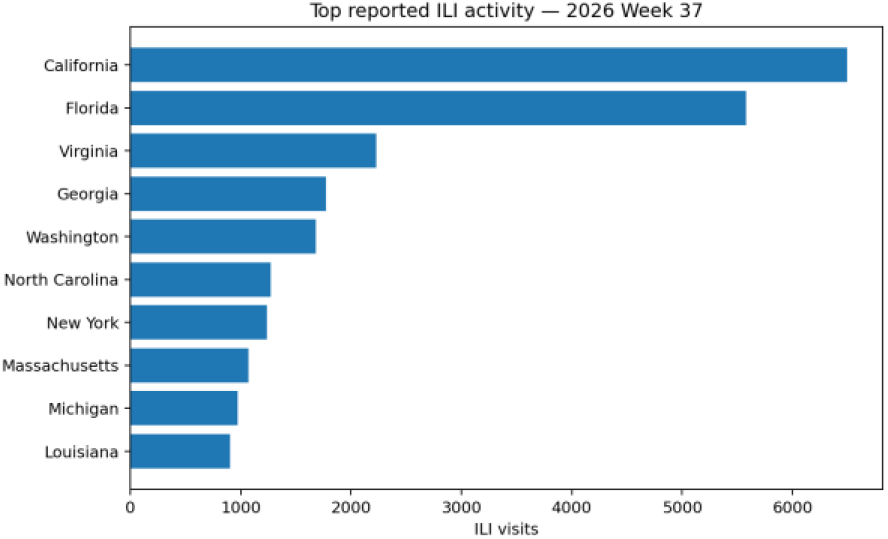
Regions with the highest reported ILI visit counts in Week 37, 2026.

It is critical to understand the difference between the amount of raw volume and statistical rarity. A location with many ILI visits might still be considered normal if its trend for that week is consistent with its past seasonal behavior. On the other hand, there can be a statistic as well when the number turns out to be relatively low in absolute terms but still varied when comparing a certain region to the given week. This is the essence of the purpose of the aberration detection process itself [5], [8], [11].

## XIV. DISCUSSION

The main finding from this framework shows that a flexible statistical pipeline is capable of transforming accurate ILINet weekly reports into reproducible week-region surveillance signals. The scope of the framework is deliberately limited compared to an epidemic detection claim: it detects deviations from historical rates of ILI, leaving the epidemiological verification of the fact to a human expert.

CUSUM produces significantly more signals than EWMA while using the same parameters. The difference should be treated as a configuration-determined difference, where CUSUM accumulates evidence over time while EWMA smooths the most recent standardized deviations using a memory parameter. Literature on the comparison of surveillance algorithms shows that results can vary according to the signals’ characteristics and the thresholds used.

The importance of the seasonal baseline cannot be understated. When a rolling baseline was used, 3314 alerts were detected in total, while only 2820 alarms were found using the seasonal reference. In the context of surveillance, the comparison of a winter week is done with just the preceding low-activity weeks, which creates an effect of unusualness. Seasonable reference, however, compares the current week with the same week in previous years.

The dashboard reveals both raw measures of surveillance and statistical measures. All surveillance parameters can be examined, from ILITOTAL, TOTAL PATIENTS, ILI rate, z-score, baseline mean, and standard deviation, CUSUM and EWMA to the final status.

This approach is in line with the ideas in the surveillance literature, where the effectiveness of the detector largely depends on baseline selection, the nature of the data being used, setting of the threshold, and characteristics of the monitored signal.

## XV. LIMITATIONS

- ILINet is synonymous with syndromic surveillance. ILI is not influenza confirmed through laboratory tests, meaning that ILI may be caused by several types of infectious agents known to affect the respiratory system [1].
- ILINet is dependent on the engagement of certain health care service providers. As reported by the CDC, the data from ILINet do not necessarily correspond to the actual number of patients in all age groups and in different places [1].
- The research does not include a sample of outbreak truth database. Hence, the values for sensitivity, specificity, positive predictive value, and false positives rate are not provided for confirmed outbreaks since to measure these characteristics, it is necessary to have certain reference events and controlled outbreaks [9], [15].
- The detector parameters are actually parameters of the project. They do not correspond to the CDC outbreak thresholds and should be validated before they are used in practice.
- The data for 2026 that are here provided are partial until week 37, thus annual comparisons including 2026 cannot be compared wholly to prior years.
- The X value is taken to be unavailable, as the exported project data does not allow for machine-code level interpretation of the character, which would preclude some false-zero entries, but would mean less number of detector inputs.
- The model applies both seasonal reference specific to a region and it does not include positivity, hospitalization, death rates, vaccination, weather, the mobility of people etc.

## XVI. CONCLUSION

EpiAlert proposes a usable and clear technique for statistical forecasting of atypical influenza-like disease monitoring flows based on actual CDC FluView ILINet data. The process can be divided into different stages of preprocessing, seasonal reference creation, CUSUM, EWMA, results assessment, and visualization. The analysis has been performed on 45,138 weekly-regional observations where 44,297 observations were valid. In total, the chosen set of parameters entered 2,820 alerts into the system. Overall, the last week of analysis generated one CUSUM signal in the state of West Virginia.

The major advantage of the method is its transparency. Every alert can be tied back to a specific regional observation, seasonal reference, standardized deviation, and two independently computed monitoring statistics.

## Data Availability

All data produced are available online at https://gis.cdc.gov/grasp/fluview/fluportaldashboard.html

## ACKNOWLEDGEMENT

The authors would like to express their sincere gratitude to Dr. Nandini K, Department of Computer Science and Engineering, Dayananda Sagar University, Bengaluru, for her valuable guidance, continuous encouragement, constructive suggestions, and supervision throughout the development of this research. The authors would also like to acknowledge the Department of Computer Science and Engineering and Dayananda Sagar University for providing the necessary resources, facilities, and academic support for this work. The authors are grateful to the faculty members and staff of the university for their guidance and encouragement throughout the project. The authors also acknowledge the Centers for Disease Control and Prevention (CDC) for making FluView ILINet influenza-like illness surveillance data publicly available, which enabled the empirical analysis and evaluation presented in this paper.

